# Rigorous Female Breast Cancer Phenotyping Using the *All of Us* Research Program

**DOI:** 10.64898/2026.08.07.26359972

**Authors:** Yuewen Qi, Kassidy Lundy-Perez, Devin Gee, Nyasha Chambwe

## Abstract

**Objectives:** Accurate phenotyping of cases and controls is essential for studying biological and environmental contributors to disease in large biobanks. We aimed to develop a flexible, customizable, and reproducible electronic health record (EHR)-based phenotyping framework for identifying disease cases and generating matched control cohorts for downstream analyses. Here, we developed the Phenotyping Algorithm for Cases and matched Controls using EHR-based Rules (PACER).

**Materials and Methods:** Applying PACER to the All of Us Research Program Curated Data Repository v8.0, we identified female breast cancer (BC) cases identified among participants recorded as female at birth using at least two BC-associated diagnostic Observational Medical Outcomes Partnership concept IDs documented at least 30 days apart. A one-to-one matched control cohort was generated by jointly matching on sex, age, genetic ancestry, and state-level residency. Clinical, socioeconomic, and genomic data were integrated for analysis.

**Results:** We identified 10,225 BC cases and generated a control cohort of the same size matched for key demographic characteristics. Comparison with a phecodeX-based BC cohort showed 91.03% agreement. Among cases responding to relevant survey items, 80.86% self-reported a personal history of BC, compared to 1.89% of controls. We detected an enrichment of BC-associated GWAS catalog variants, pathogenic mutations in known risk genes, and higher polygenic risk scores in cases compared to controls.

**Discussion and Conclusion:** Concordance across a phecodeX-based cohort, self-reported survey responses, and genomic analyses supports the validity of PACER-defined cohorts. PACER is publicly available and readily adaptable to other diseases, supporting future research in risk modeling and precision medicine.

## Introduction

Large population-scale biobanks such as the UK Biobank and the National Institutes of Health *All of Us* Research Program (*AoU*) enable the study of diverse populations and the integration of both biological and environmental factors in health-related research (1,2). *AoU*, designed to be one of the largest and most demographically diverse population databases in the United States, aims to enroll over one million participants, with particular emphasis on populations historically underrepresented in biomedical research in the United States (U.S.) (3). *AoU* provides access to an extensive data catalog that includes electronic health records (EHRs), physical measurements, wearable device data, genomic information and patient-reported surveys on topics that include baseline demographics, socioeconomic conditions, lifestyle, medical history and environmental exposures. This compendium of detailed baseline and longitudinal health-related information allows researchers to more comprehensively study how genetic and socioeconomic factors interact and influence disease risk and health outcomes across populations (4). Unlike narrowly defined cohorts designed to study a single disease, *AoU* enables researchers to design studies with a variety of focuses including population-related research and disease focused studies. The breadth of diverse disease conditions available in *AoU* makes it possible to identify risk factors, find better treatments, and advance precision medicine.

To identify the subset of biobank participants to use as cases and controls for a given disease or condition, reliable EHR-based phenotyping methods are required. However, because EHR systems were designed primarily for clinical and administrative use rather than research, EHR-based cohorts may be affected by record missingness, recording error, misclassification, and selection bias, including the underrepresentation of patients with fragmented or limited access to care (5,6). Previous studies have utilized EHR data to define case cohorts using one or more diagnostic codes, some requiring at least 2 occurrences separated by a minimum time interval, often 30 days, to reduce false-positive classification (7–11). Widely used EHR-based phenotyping algorithms include phecodeX (12) and those from the Phenotype KnowledgeBase (PheKB) by the Electronic Medical Records and Genomics (eMERGE) Network (13), which are made publicly available for the research community. Prior work evaluating three levels of complexity for phenotyping algorithms for ovarian, female breast, and colorectal cancers using an earlier release version of the *AoU* dataset showed that phenotype complexity can substantially affect statistical power in genomic association testing, and that no single phenotyping approach performs best across all cancer types (8). While larger control cohorts may improve statistical power in genomic analyses, studies that integrate genomic and environmental contributors to disease also require well-matched controls on important demographic factors, such as age and geographic location, to reduce selection bias and strengthen downstream analysis (14). Together, these findings highlight the need for a flexible framework that enables easy and efficient definition and modification to phenotyping inclusion, exclusion, and filtering criteria for case-control cohorts across different disease conditions.

Here, we developed a scalable and reproducible phenotyping framework, **P**henotyping **A**lgorithm for cases and matched **C**ontrols using **E**HR-based **R**ules (**PACER**), for constructing well-defined case and demographically matched control cohorts within *AoU* that facilitate more balanced analyses of both biological and environmental contributors to disease across multiple disease types. To demonstrate its utility, we applied PACER to defining a female breast cancer (BC) case-control cohort. BC is a common and multifactorial disease influenced by genetic, clinical, lifestyle, and socioeconomic factors (15). This complexity makes it a suitable use case for evaluating a framework that integrates EHR, genomic, and socioeconomic data. In addition, BC shows significant disparities in the United States, as Black women experience 40% higher BC mortality despite having lower incidence compared to non-Hispanic White women (16), highlighting the importance of constructing case and well-matched control cohorts when studying etiological risk factors across populations.

Using PACER, we identified an *AoU*-based BC case cohort using EHRs and constructed a one-to-one matched control cohort from a control pool we assembled by matching on sex at birth, age, predicted genetic ancestry, and state-level residency. To evaluate our approach, we cross-matched our BC case cohort with cases identified by phecodeX (12), examined agreement with self-reported BC status from participant survey data not used in the phenotyping framework, and calculated the prevalence of known BC risk variants and the distribution of genetic risk between cases and controls. Our results demonstrate the validity of our approach while highlighting study-specific nuances needed for a more customizable cohort phenotyping approach with flexible matching strategies for generating case and matched control cohorts across disease types. This framework is fully customizable and can be extended to other disease studies within population biobanks, allowing researchers to adapt and compare different cohort definitions and matching strategies to meet their study objectives.

## Materials and Methods

The algorithmic logic of PACER is outlined in (**Figure 1**) and demonstrated here using participant data from the *AoU* Curated Data Repository (CDR Version 8.0 Controlled Tier) dataset to identify a matched BC case-control cohort.

**Figure 1.**
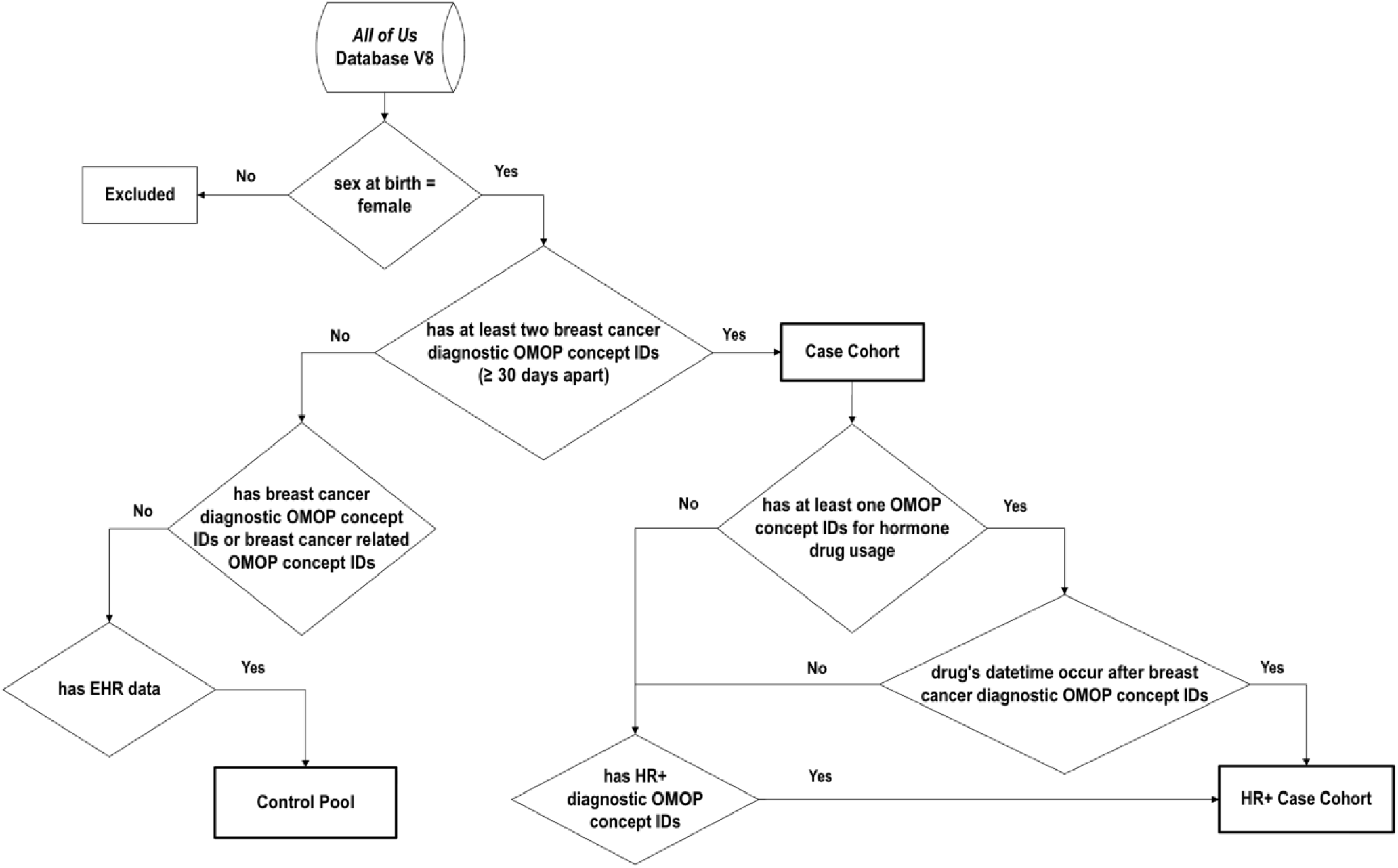
PACER Breast Cancer Phenotyping Workflow. The overall pipeline for identifying a breast cancer (BC) case cohort, hormone receptor positive (HR+) case cohort, and a pool of controls using the *All of Us* (*AoU)* V8 dataset. Only participants with “female sex at birth” are included. Participants with at least 2 BC diagnostic OMOP concept IDs occurring at least 30 days apart are classified as cases. BC cases with HR+ diagnostic OMOP concept IDs or OMOP concept IDs for hormone drug usage with a “datetime” stamp after BC diagnostic OMOP concept IDs are classified as HR+ cases. Participants without any BC diagnostic OMOP concept IDs or BC-related OMOP concept IDs are included in the control pool.

### PACER case cohort determination

We classified participants as BC cases if they were female sex at birth (“sex_at_birth”) and had at least 2 BC diagnostic Observational Medical Outcomes Partnership (OMOP) concept IDs documented at least 30 days apart. This approach builds on methods and OMOP concept IDs from Ning et. al.(17), and includes additional BC OMOP concept IDs identified in the *AoU* data dictionary (**Table S1**). We further subclassified the BC case cohort into hormone receptor-positive (HR+) participants due to their distinct biology and methods for treatment. Participants were classified into BC HR+ case cohort if they satisfied all criteria for BC case classification and either had diagnostic OMOP concept IDs indicating HR+, estrogen receptor positivity (ER+), or progesterone receptor positivity (PR+) (**Table S2**), or had records indicating relevant hormone drug use (**Table S3**) after the date of their BC diagnostic OMOP concept IDs.

### PACER control pool selection

We defined a BC control pool of all *AoU* participants that met the following criteria: had EHR data available, was recorded female sex at birth (“sex_at_birth”), had no BC diagnostic OMOP concept IDs (**Table S1**), and no BC-related OMOP concept IDs that indicate history or possibility of BC (**Table S4**). This represents the base control cohort if no additional matching criteria are required depending on study objectives.

### PACER control cohort matching strategy

To generate a matched control cohort suitable for integrative downstream genomic, clinical, and socioeconomic analyses, we selected samples from the control pool by applying matching criteria to the case cohort. First, sex at birth was matched between the case and control cohort. Second, participant age was calculated using participants’ date of birth and the present date. Controls were matched across the age range represented in the case cohort, binned in 10-year increments. Third, we incorporated *AoU*-provided genetic ancestry estimated from short-read whole genome sequencing (srWGS) into the matching process. Genetic ancestry was inferred using the outputs of the random forest classifier model run by AOU (18). Briefly, 16 principal components were calculated from genotype data from the Human Genome Diversity Project and 1000 Genomes samples. Participants were assigned to a predicted genetic ancestry group if the probability of group classification was at least 0.75; participants without a dominant ancestry group were labeled “OTH,” and those without srWGS data were labeled “no_ancestry.” Matching was performed proportionally across all ancestry categories: AFR, AMR, EAS, EUR, MID, OTH, no_ancestry, and SAS (**Table 1**). Fourth, state of residence was derived by mapping participants’ three-digit ZIP codes to U.S. states. The control cohort was selected by randomly choosing participants while jointly matching on sex at birth, age, predicted ancestry, and state residency rather than matching each variable independently. We set the control cohort size to a one-to-one ratio with the case cohort. However, the matching algorithm is flexible and allows researchers to customize cohort size as needed.

**Table 1.** Demographic summary of the PACER BC case and matched control cohorts. *P-values are calculated using the Mann-Whitney U test (MWU) between the case and the matched control cohorts for numerical factors such as age; and by the chi-square test for categorical factors such as race, ethnicity and predicted genetic ancestry.

| Factors | Category | Case (n)(%) | Control (n)(%) | *P value |
| --- | --- | --- | --- | --- |
| **N** |  | 10225(100.00) | 10225(100.00) |  |
| **Sex at Birth** | Female | 10225 (100.00) | 10225 (100.00) | — |
| **Age, y** | <40 | 114 (1.12) | 129 (1.26) | 0.497 |
|  | 40–49 | 613 (6.00) | 620 (6.06) |  |
|  | 50–59 | 1485 (14.52) | 1492 (14.59) |  |
|  | 60–64 | 1212 (11.85) | 1214 (11.87) |  |
|  | ≥65 | 6801 (66.51) | 6770 (66.21) |  |
| **Race** | White | 7040 (68.85) | 6753 (66.04) | <0.001 |
|  | Black/Afr. Am. | 1254 (12.26) | 1222 (11.95) |  |
|  | Others/ Haw./Pac. Isl. | 1083 (10.59) | 1199 (11.72) |  |
|  | Multiracial | 312 (3.05) | 322 (3.15) |  |
|  | Asian | 230 (2.25) | 220 (2.15) |  |
|  | Am. Indian/Alaska Nat. | 63 (0.62) | 81 (0.79) |  |
|  | Mid. East/N. Afr. | 55 (0.54) | 68 (0.67) |  |
|  | Skipped | 188 (1.84) | 360 (3.52) |  |
| **Ethnicity** | Non-Hispanic | 8779 (85.86) | 8508 (83.21) | <0.001 |
|  | Hispanic | 1166 (11.40) | 1197 (11.71) |  |
|  | Other | 92 (0.90) | 160 (1.56) |  |
|  | Skipped | 188 (1.84) | 360 (3.52) |  |
| **Genomic Data (srWGS)** | Yes | 8576 (83.87) | 8574 (83.85) | 0.985 |
|  | No | 1649 (16.13) | 1651 (16.15) |  |
| **Predicted Genetic Ancestry (Probability ≥ 0.75)** | EUR | 5891 (57.61) | 5909 (57.79) | 0.456 |
|  | AFR | 1162 (11.36) | 1166 (11.40) |  |
|  | AMR | 781 (7.64) | 784 (7.67) |  |
|  | MID/OTH | 551 (5.39) | 544 (5.32) |  |
|  | EAS | 155 (1.52) | 146 (1.43) |  |
|  | SAS | 36 (0.35) | 25 (0.24) |  |
|  | No ancestry data | 1649 (16.13) | 1651 (16.15) |  |

### PhecodeX-based case cohort determination

An PhecodeX phenotyping was performed using the “createPhenotypes()” function of the PheWas R package (version 0.99.6-1). *AoU* participants’ condition codes were mapped to their respective phecode using the provided phecode map. BC cases were defined as participants that had 2 condition codes that were 30 days apart for the female specific BC phecode label “CA_105.1”.

### Self-Identified case cohort determination

We defined a BC case cohort based on participants who self-reported BC by selecting “self” in response to the question “Including yourself, who in your family has had breast cancer?” from the “Personal and Family Health History” survey.

### Demographic, clinical and socioeconomic variable abstraction

Potential study-relevant participant variables such as basic cohort demographic information, clinical measures such as BMI and self-reported lifestyle and socioeconomic status indicators were abstracted from collected EHR and self-reported survey data across the case-control cohort (**Table 1**; **Table 2; Table S5**). Demographic information including “sex-at-birth”, “education level”, “employment status”, “income”, “insurance (Yes/No)”, “insurance type” were extracted from “the Basics,” survey. BMI values were extracted from the *AoU* Measurement Domain using OMOP concept IDs 4245997, 3038553, and 44783982. BMI was classified as Underweight (BMI < 18.5), Normal weight (18.5 ≤ BMI < 25), Overweight (25 ≤ BMI < 30), and Obesity (BMI ≥ 30) following the World Health Organization definition. Participants without available BMI data were labeled as “No data”. Individual level lifestyle variables such as ‘alcohol use’ and ‘smoking frequency’ were extracted from the “Lifestyle” survey. An area-level socioeconomic status indicator, ‘deprivation index’ was extracted from *AoU* “Zip Code Socioeconomic Status Data” in *AoU* workbench. The deprivation index is a composite metric derived by *AoU* using principal components analysis of six U.S. Census-based measures from the 2015 American Community Survey (ACS) and is calculated by *AoU* using only the first three digits of participants’ ZIP codes, a privacy-preserving approach that limits geographic specificity by mapping participants to broad regions rather than precise neighborhoods.

**Table 2.** Socioeconomic and clinical summary for the PACER BC case and matched control cohort. *P-values are calculated using the Mann-Whitney U test (MWU) between case and control cohorts for numerical factors such as BMI and area deprivation index; and by the chi-square test for categorical factors such as education level, employment status, income, insurance, insurance type, alcohol use (drink frequency), and smoking frequency.

| Factors | Category | Case (n)(%) | Control (n)(%) | *P value |
| --- | --- | --- | --- | --- |
| **N** |  | 10225(100.00) | 10225(100.00) |  |
| **BMI** | Underweight | 203 (1.99) | 242 (2.37) | 0.007 |
|  | Normal weight | 2983 (29.17) | 2874 (28.11) |  |
|  | Overweight | 3056 (29.89) | 2854 (27.91) |  |
|  | Obesity | 3939 (38.52) | 4065 (39.76) |  |
|  | No data | 44 (0.43) | 190 (1.86) |  |
| **Area Deprivation Index** | Mean | 0.313 | 0.311 | 0.024 |
|  | Median | 0.302 | 0.302 |  |
| **Education Level** | Advanced degree | 2901 (28.37) | 2621 (25.63) | <0.001 |
|  | College graduate | 2724 (26.64) | 2458 (24.04) |  |
|  | College 1–3 | 2676 (26.17) | 2670 (26.11) |  |
|  | 12 / GED | 1273 (12.45) | 1511 (14.78) |  |
|  | Grade 9–11 | 251 (2.45) | 417 (4.08) |  |
|  | Grade 5–8 | 152 (1.49) | 201 (1.97) |  |
|  | ≤Grade 4 | 89 (0.87) | 129 (1.26) |  |
|  | Skipped | 159 (1.56) | 218 (2.13) |  |
| **Employment Status** | Retired | 4292 (41.98) | 4327 (42.32) | <0.001 |
|  | Employed | 3114 (30.45) | 2801 (27.39) |  |
|  | Unable to work | 986 (9.64) | 1067 (10.44) |  |
|  | Self-employed | 648 (6.34) | 587 (5.74) |  |
|  | Homemaker | 517 (5.06) | 543 (5.31) |  |
|  | Unemployed >1 year | 308 (3.01) | 397 (3.88) |  |
|  | Student/Skipped | 213 (2.08) | 285 (2.79) |  |
|  | Unemployed <1 year | 147 (1.44) | 218 (2.13) |  |
| **Income** | >200k | 916 (8.96) | 646 (6.32) | <0.001 |
|  | 150k–200k | 626 (6.12) | 547 (5.35) |  |
|  | 100k–150k | 1286 (12.58) | 1108 (10.84) |  |
|  | 75k–100k | 1094 (10.70) | 967 (9.46) |  |
|  | 50k–75k | 1248 (12.21) | 1243 (12.16) |  |
|  | 35k–50k | 849 (8.30) | 867 (8.48) |  |
|  | 25k–35k | 683 (6.68) | 744 (7.28) |  |
|  | 10k–25k | 977 (9.56) | 1205 (11.78) |  |
|  | <10k | 546 (5.34) | 842 (8.23) |  |
|  | Skipped | 2000 (19.56) | 2056 (20.11) |  |
| **Insurance (Yes/No)** | Yes | 9942 (97.23) | 9750 (95.35) | <0.001 |
|  | No | 114 (1.11) | 273 (2.67) |  |
|  | Do not know/Skipped | 169 (1.66) | 202 (1.97) |  |
| **Insurance Type** | Medigap/Medicare | 2180 (21.32) | 2137 (20.90) | <0.001 |
|  | Employer/Union | 1763 (17.24) | 1515 (14.82) |  |
|  | Medicaid | 661 (6.46) | 836 (8.18) |  |
|  | Purchased | 505 (4.94) | 446 (4.36) |  |
|  | Private | 430 (4.21) | 347 (3.39) |  |
|  | Other health plan | 128 (1.25) | 130 (1.27) |  |
|  | Military/VA | 113 (1.10) | 106 (1.03) |  |
|  | Other Govt/State sponsored | 44 (0.43) | 53 (0.52) |  |
|  | Single service/ SCHIP/Indian/No coverage | 33 (0.32) | 23 (0.23) |  |
|  | Do not know/Skipped | 4368 (42.72) | 4632 (45.30) |  |
| **Alcohol Use** | ≥4 per week | 1222 (11.95) | 1181 (11.55) | 0.086 |
|  | 2–3 per week | 1280 (12.52) | 1147 (11.22) |  |
|  | 2–4 per month | 1774 (17.35) | 1717 (16.79) |  |
|  | Monthly or less | 3100 (30.32) | 3092 (30.24) |  |
|  | Never | 1829 (17.89) | 1850 (18.09) |  |
|  | Skipped | 118 (1.15) | 144 (1.41) |  |
| **Smoking Frequency** | Not at all | 3262 (31.90) | 3128 (30.59) | <0.001 |
|  | Some day | 260 (2.54) | 316 (3.09) |  |
|  | Every day | 408 (3.99) | 718 (7.02) |  |
|  | Do not know/Skipped | 46 (0.45) | 47 (0.46) |  |

### Genetic predisposition analyses

For all genomic analyses, we considered cases and the full control pool (no strict matching) as ‘controls’ for all participants with srWGS data availability (8,576 cases, 131,857 controls). We analyzed common variants that were pre-filtered by *AoU* population-specific allele frequency > 1% or population-specific allele count > 100 using PLINK version 1.9 (18,19). The following variant quality filters were applied: call rate > 0.9, Hardy-Weinberg equilibrium (p value threshold = 10^−15^), and excess heterozygosity-associated thresholds (p-value threshold = 10^−15^).

#### Disease associated variant analysis

Disease trait variant associations were downloaded from the NHGRI-EBI Catalog of human genome-wide association studies (GWAS catalog) (19). GWAS associations were subset to keep SNPs associated with BC and then analyzed for overlap with variants in our cohort by genomic position. We used the absolute count of GWAS BC-associated variants to split participants into high burden (above median) and low burden (below median) groups. Fisher’s exact test determined the enrichment of high burden participants versus low burden participants between PACER cases and controls.

#### Pathogenic variant prevalence analysis

We curated a list of 14 genes implicated in BC predisposition (*BARD1, BRCA1, BRCA2, BRIP1, CDH1, CHEK2, NF1, PALB2, RAD50, RAD51C, RAD51D, RB1, STK11, TP53*) (20,21). Using the *AoU* curated ClinVar variant callset, we subset variants within these 14 predisposition genes and filtered the set to keep variants annotated as pathogenic or likely pathogenic with no conflicting reports. We used Fisher’s exact test to calculate odds ratios of pathogenic variant enrichment between cases and controls for each individual gene across the full cohort and for groups defined by primarily African or European ancestry.

#### Polygenic Risk Scoring

We calculated a BC associated polygenic risk score (PRS) in our case control cohort using published variant weights (PGS004688) (20) from the PGS Catalog (21).

## Results

### Cohort characteristics

Using PACER (**Figure 1**, detailed inclusion criteria outlined in **Methods**), we identified 10,225 BC cases from all *AoU* participants (*AoU* CDR v8.0). Of these BC cases, 8,576 (83.87%) had srWGS and predicted genetic ancestry available (**Table 1**) and 5,386 (52.67%) were further subtyped as HR+ BC cases, of which 4,555 (84.57%) have srWGS data available (**Table S5**). We identified 166,987 participants for the control pool, from which we generated an age, genetic ancestry-distribution and state-level residence matched control cohort of 10,225 participants, of whom 8,574 (83.85%) had srWGS data available (**Table 1**).

We developed a fully automated pipeline to generate data for cohort-level summary tables for potential key study demographic, lifestyle, risk factor and socioeconomic status indicators using data from self-reported participant surveys (**Table 1**; **Table 2; Table S5**). This pipeline can be applied to other custom-defined *AoU* cohorts and modified to fit a given study’s objectives. We examined differences in clinical and socioeconomic characteristics between BC cases and controls and found modest shifts toward higher educational attainment, household income and insurance coverage in cases compared to controls (**Table 2**, Chi-square test p < 0.001). BC cases tended to have Medigap/Medicare, employer-sponsored medical or private insurance compared to higher Medicaid coverage in the controls (**Table 2**, Chi-square test p < 0.001). The BC case cohort had a higher proportion of overweight participants compared to controls, in agreement with the established relationship between BMI and BC risk (22). These demographic, lifestyle, and socioeconomic factors present a summary of the selected cohort along key study variables of interest and can be customized for each study.

### Control cohort matching criteria minimizes potential bias in participant selection

The rationale for applying control cohort matching criteria depends on a particular study’s objectives. For example, in this instance, we observed that the age distribution of the BC case cohort and the full control pool differed substantially, with the case cohort skewing towards older age, while the control pool showed a bimodal distribution with peaks around 35 and 63 years old (**Figure 2A**, Mann-Whitney U test, p < 0.001). This discrepancy is not unexpected, as age is a known risk factor for BC (23), and the control pool represents a subset of the *AoU* population that can be ruled out as having any form of cancer, but is generally sampled from the adult US population. Therefore, an age distribution constraint on the selection of control participants as shown in **Figure 2B** is necessary to eliminate confounding effects attributed to aging rather than the disease condition under study.

**Figure 2.**
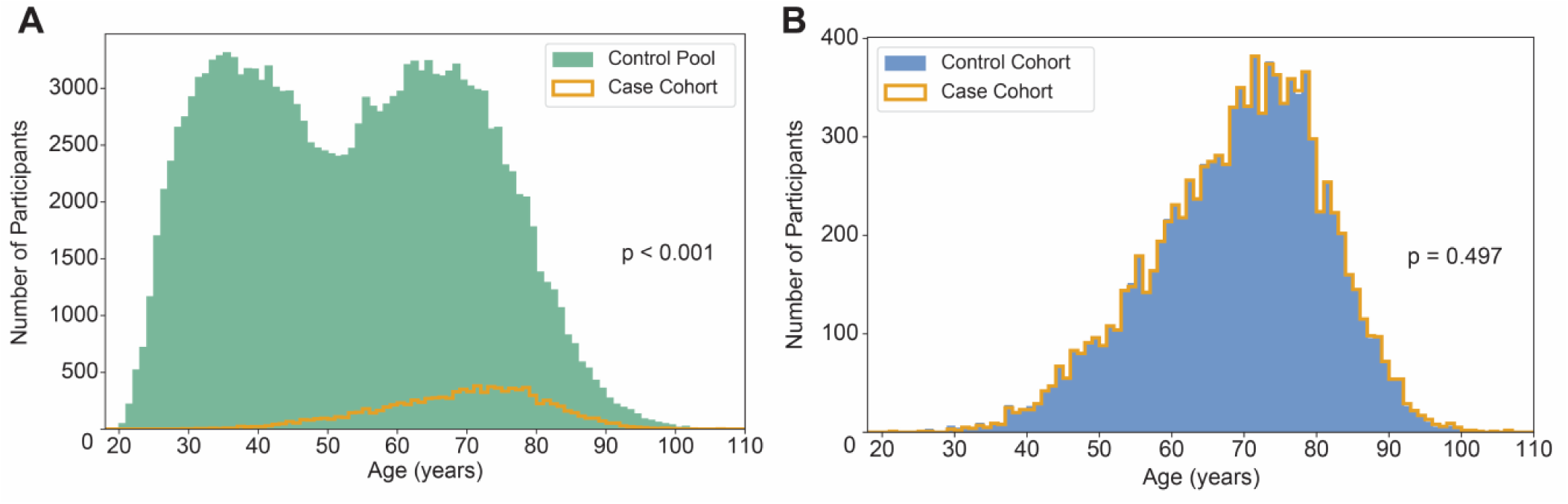
Comparative analysis of PACER cohort age distributions. (A) Histogram depicting the control pool age distribution (green shading) overlaid with the case cohort age distribution (orange line). The distributions differ significantly (p < 0.001, Mann–Whitney U test), demonstrating the need for age matching between case and control cohorts. (B) Histogram showing the age distribution in the 1:1 sex at birth, state-level geographic location and age-matched control (blue shading) and case cohort (orange line). The similarity between the two distributions shows effective age matching (p = 0.497, Mann– Whitney U test).

In other instances, a study with the objective of assessing the impact of genetic variation on disease could be confounded by underlying population-specific effects. The racial and ethnic diversity and population migration history of the U.S. magnifies these effects. The availability of srWGS across much of the *AoU* population (∼83-84% in this cohort) allows for the inference of genetic ancestry based on reference population that allows for case-control cohort matching based on ancestry. Here we show that predicted genetic ancestry distributions were consistent between cases and controls after matching. Principal component analysis of srWGS genotype calls revealed that the case and control cohorts had similar patterns of clustering and admixture across the 6 reference genetic ancestry groups (**Figure S1**), and the control cohort also showed a similar distribution of ancestry assignment probabilities (**Figure 3A-B**). Although race and ethnicity were not explicitly used for matching, their distributions were broadly similar across cohorts (**Figure 3 B-C**; **Table 1**), which may partly reflect their non-random overlap with genetic ancestry variables used in matching.

**Figure 3:**
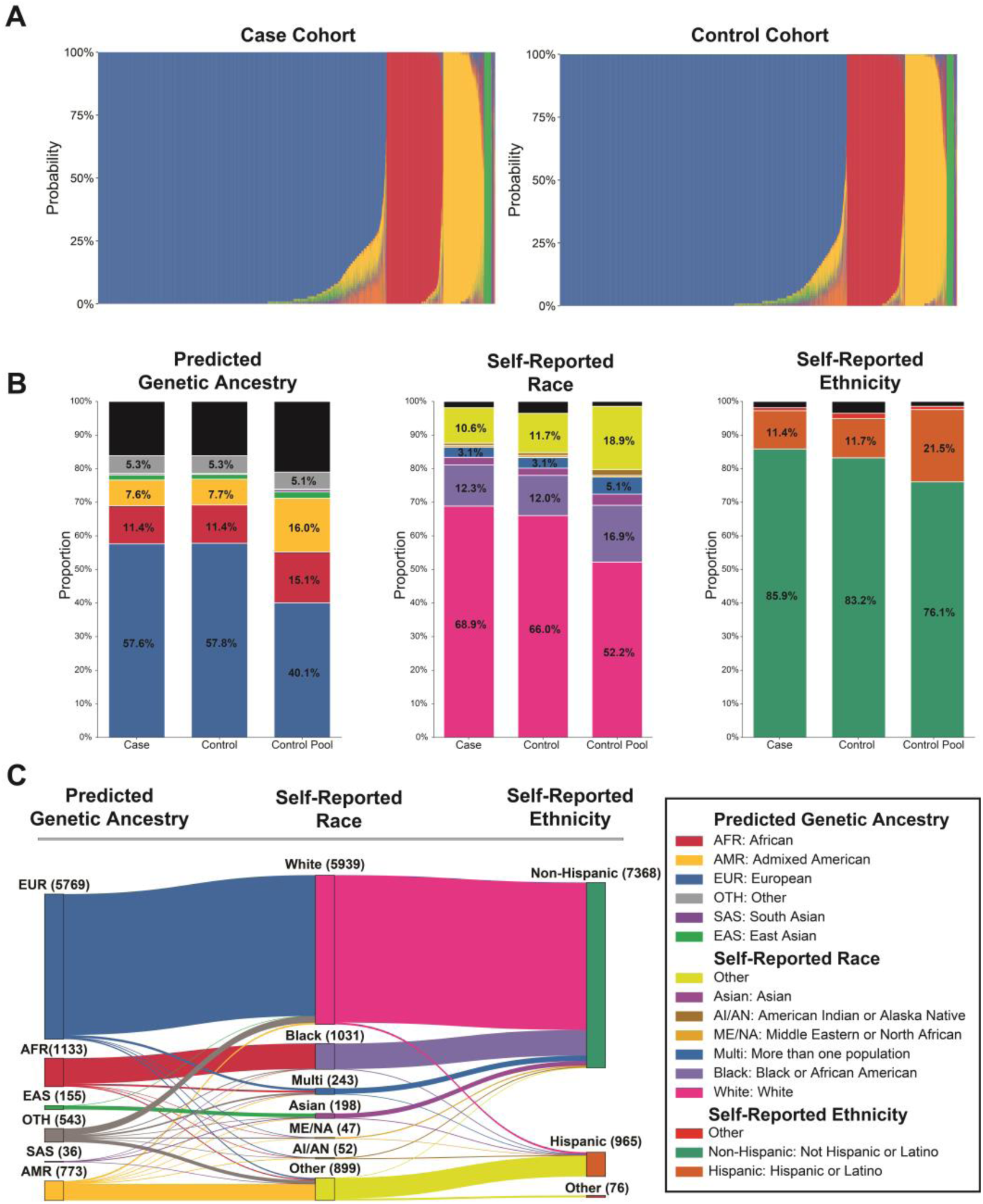
Genetic ancestry, race, and ethnicity distributions for the PACER BC case and control cohorts. (A) Stacked bar plots showing the probability of being assigned to each of the six reference genetic ancestry groups (y-axis) for each individual participant with predicted genetic ancestry data (x-axis) in each cohort (B) Proportion of cases for each cohort in each predicted genetic ancestry group (left), self-reported race (middle), and self-reported ethnicity (right) class. (C) An alluvial plot illustrating the relationships between ethnicity, race, and predicted genetic ancestry for cases with complete data across all three variables.

Additionally, the *AoU* population is sampled from across the U.S. with a concentration of study participants from states with larger population sizes and *AoU*-funded enrollment centers. The geographic distribution of BC cases across U.S. states in the *AoU* dataset shows that California, Pennsylvania, and Massachusetts recruited the largest numbers of BC cases, likely reflecting the greater numbers of *AoU* participants with available EHR data within *AoU* in these states (**Figure 4A).** In most states, the state-level case-to-control ratio is close to 1:1 **(Figure 4B)**, consistent with our state-level matching strategy. Deviations were most apparent in states with smaller case counts, where small differences in eligible controls shifted the ratio. For example, Vermont and Delaware had relatively higher case-to-control ratios, whereas Utah, South Dakota, and Arkansas had lower ratios. These differences reflect both underlying sample sizes and the matching priority assigned to age and ancestry over state residency when exact matches were unavailable. Despite these minor variations, state-level matching may have reduced the risk of geographic sampling bias for downstream analyses.

**Figure 4.**
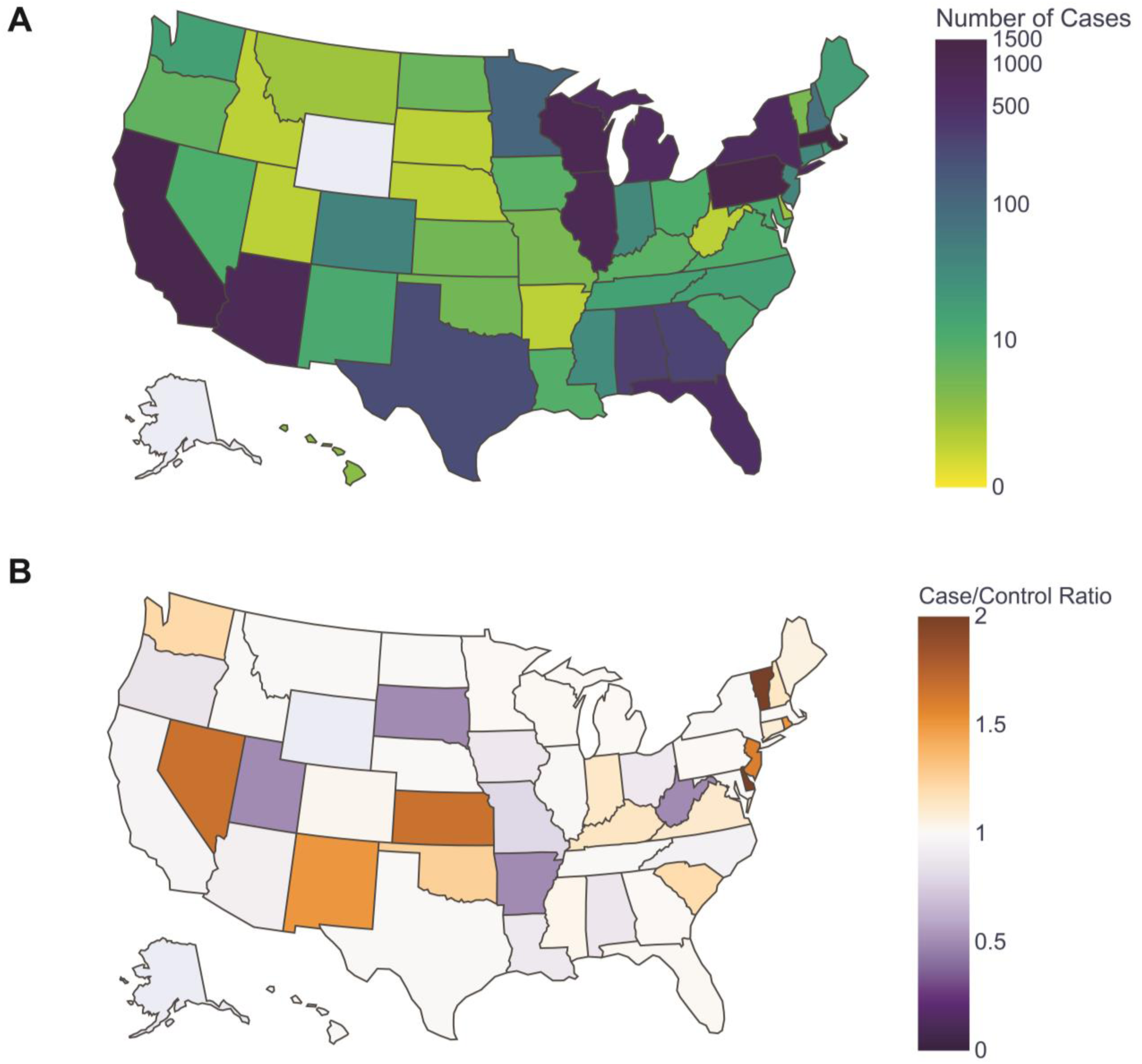
State-level geographic distribution of the PACER BC case and matched control cohort. (A) The distribution of the number of BC cases in the PACER-defined BC case cohort across U.S. states. Color scale indicates the number of BC cases with deeper blue indicating a higher case count and deeper yellow indicating a lower case count in a given state. (B) The relative sampling distribution of BC cases and controls for each U.S. states. States are colored by the case-to-control ratio, with deeper orange indicating a higher proportion of cases and purple indicating a higher proportion of controls. States with no participants in the BC case cohort (Wyoming and Alaska) are shown in gray.

### PACER concordance with phecodeX and BC self-report

To assess concordance in phenotyping with independent approaches, we evaluated case ascertainment agreement with self-reported survey data and phecodeX (12). Within the case cohort, 6,342 participants responded to at least one survey item, and 5,128 self-reported as having BC themselves, corresponding to a rate of 80.86% based on survey data independent of EHR. In the matched control cohort, 5,384 participants answered at least one survey question, of whom 102 (1.89%) self-reported having BC. This substantial difference in self-reported BC status between the case and control cohorts demonstrates consistency between PACER-generated cohorts and survey data.

When comparing the PACER-derived BC case cohort to the one generated by phecodeX (12), we found 91.03% overlap between the two with 4.73% of cases exclusive to PACER and 4.24% of cases exclusive to phecodeX (**Figure S2**). Like PACER, 80.97% of phecodeX cases are self-reported as having BC. Differences between the retrieved cohorts emerged from PACER’s more expansive list of BC OMOP concept IDs (**Table S1**) compared to phecodeX’s inclusion of an observation history of BC. There were no significant differences in the distribution of racial or ethnic groups between the cohorts. PACER achieves similar performance to phecodeX in phenotyping BC while providing additional options for customization and the ability to select match controls based on study-specific criteria.

### Genomic Validation of the PACER-Defined Case-Control Cohort

To demonstrate the robustness of PACER for identifying high-fidelity BC cases, we performed genomic analyses on 140,433 participants with srWGS data available (8,576 cases, 131,857 control pool). The underlying rationale for these analyses is that *bona fide* BC cases should be enriched for known BC genetic risk associations, particularly in well-established BC predisposition genes, and should exhibit a higher overall BC-associated polygenic risk profile.

First, we compared the prevalence of known BC risk alleles between cases and controls by identifying variants in our cohort that overlapped with published BC-associated GWAS SNPs from the GWAS catalog (24). Of the 3,182 BC-associated SNPs in the GWAS catalog, 1,789 met the genome-wide significance threshold (p < 5 ⋅ 10^−8^) for BC association across the PACER determined case control cohort **(Figure S3)**. Cataloged risk alleles were significantly enriched in the case cohort for participants of both African (AFR p = 0.006) and European ancestry (EUR p < 0.001) (**Figure 5A**). Enrichment and risk allele burden were both higher in EUR participants, which can largely be explained by the well-documented underrepresentation of non-European subjects in the GWAS catalog (25).

**Figure 5:**
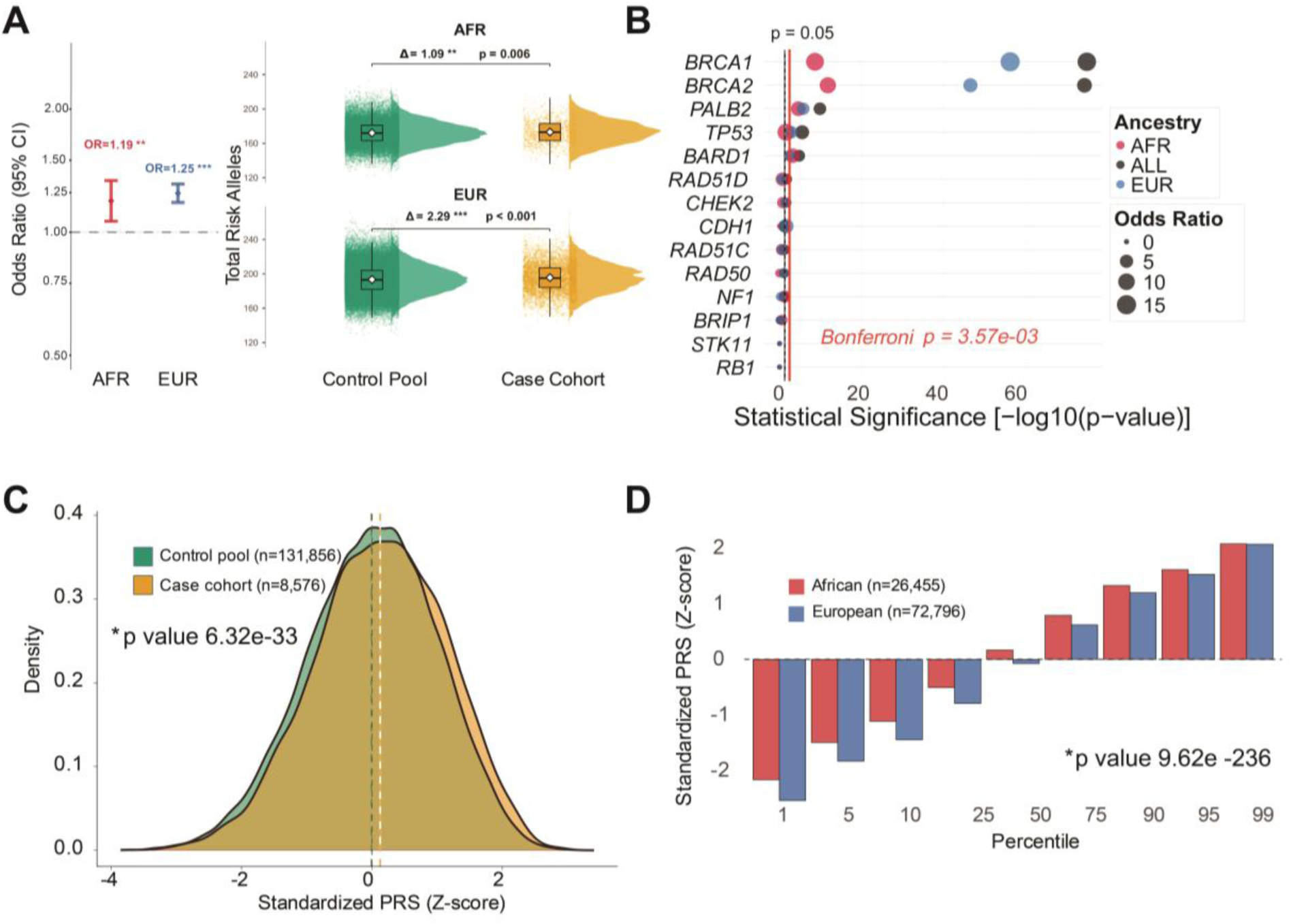
Genetic predisposition analyses in the PACER BC case control cohort. (A) (Left) shows case, control and ancestry-specific context provided for the 1,789 overlaps in cases with both position and genotype overlap. Fisher’s exact test and a median-split approach are used to identify participants as high burden (above the median) or low burden (below the median). Odds ratios (ORs) show enrichment of high burden participants in cases compared to controls across ancestries. (Right) Absolute counts of risk alleles are shown across ancestries. (B) shows ORs of pathogenic and likely-pathogenic (PLP) variant carriers of variants within BC-associated predisposition loci in cases versus controls. Fisher’s exact test is used. (C) shows polygenic risk score assessment of participants in the case cohort and control pool. (D) shows PRS values by percentile among participants with African and European predicted genetic ancestry.

Second, we analyzed the enrichment of ClinVar-annotated pathogenic or likely pathogenic (P/LP) variants in high and moderate penetrance breast cancer susceptibility genes (26,27). *BRCA1, BRCA2 and PALB2* showed statistically significant differences in P/LP variant carrier frequency between cases and controls at a conservative significance threshold (Bonferroni corrected p = 3.57×10^-03^) (**Figure 5B**). *TP53* reached significance in EUR ancestry subset but not in the AFR subset suggesting that the cohort may be underpowered to detect effects in this subgroup. Not strong evidence of genetic predisposition was found across the remaining genes tested.

Third, we calculated polygenic risk scores (PRS) in the PACER BC case control cohort using published variant weights (PGS004688 (22)) from the PGS Catalog (21). As expected, the mean PRS was significantly higher in cases than in controls, though with modest effect sizes (p = 6.83 ⋅ 10^−3^ and Cohen’s coefficient 0.134) (**Figure 5C**), suggesting that the PRS only partially captures the full BC risk spectrum. PRS distributions stratified by percentile and ancestry revealed significantly different risk scores between AFR and EUR ancestry BC cases (p = 9.62 ⋅ 10^−236^) (**Figure 5D**).

Overall, these genomic analyses demonstrate that the PACER-defined cohort largely captures the expected genomic signals, with significant enrichment of BC-associated variants in the case cohort and consistent evidence of ancestry-specific patterns. While publicly available PRS scores explain a portion of the observed risk, the modest effect sizes underscore the need for careful interpretation when PRS models are applied to diverse populations.

## Discussion

In this study, we demonstrated the utility and feasibility of the PACER framework applied to *AoU* program data to construct large BC case and matched control cohorts for downstream analyses. *AoU* provides a uniquely rich resource for integrating EHR, genomic, and socioeconomic data, allowing for comprehensive investigation of disease risk factors once well-defined cohorts are established (10). PACER also generates a summary of genomic, clinical and socioeconomic variables, enabling rapid evaluation and comparison of different phenotyping strategies and making it straightforward to test and refine cohort definitions. The full algorithm is available on a community workspace within the *AoU* Researcher Workbench, allowing researchers to reproduce and adapt the pipeline across diseases. A central component of the PACER framework is the construction of a matched control cohort from a defined control pool rather than using all other available participants as controls. Specifically, we constructed a matched control cohort that closely reflects the distribution of the case cohort on sex at birth, age, predicted genetic ancestry, and state-level residency. This is particularly important for studies investigating both genomic and environmental contributors to disease, where differences in demographic or geographic distributions can introduce bias (14).

We evaluated the PACER framework using three independent orthogonal approaches not used in cohort construction. Specifically, ∼90% of participants defined in the PACER case cohort overlapped with BC cases identified by the phecodeX algorithm. In addition, 81% of case participants self-reported a personal history of BC in the *AoU* “Personal and Family Health History” survey, compared to ∼2% in the control cohort. Finally, we observed expected patterns in the distribution of variants associated with BC, providing additional biological support for the validity of the PACER-identified cohorts. Together, these findings indicate that PACER captures consistent and concordant signals across clinical records, self-reported survey responses, and genomic data, supporting the validity of the framework.

A key strength of PACER is in its flexibility to adapt to various phenotyping definitions. Prior work has shown that phenotype definitions can greatly influence statistical power in genomic analysis, and that no single approach fits all diseases (8). PACER provides a structured, flexible, and reproducible framework that allows researchers to define and modify phenotyping and matching criteria as needed through inclusion and exclusion criteria. Specifically, researchers can vary key components of cohort construction, including the choice of diagnostic OMOP concept IDs, the number of required OMOP concept ID occurrences and the time interval between OMOP concept IDs. Control cohorts can be generated using adjustable matching strategies from a control pool, enabling systematic comparison of cohorts based on the same dataset. Matching criteria can be included, relaxed, or omitted depending on the research question. This is particularly important in large biobank settings, where overly restrictive criteria can limit sample size, and overly broad definitions can introduce misclassification. We do note however, that joint matching can produce edge cases in which no participants in the control pool can be matched to a given case based on the full combination of selected matching criteria. To address this, the PACER algorithm defines matching priorities. For this study, sex at birth was assigned the highest matching priority because the BC case cohort included only participants self-reported as female at birth, and the control pool was restricted to the same category. However, when multiple sex at birth categories are included in the case cohort, PACER preserves their corresponding proportions in the matched control cohort. Age and predicted ancestry were also prioritized and matched exactly unless no exact match or combination of matching criteria was available. State residency was treated as a lower-priority criterion and was relaxed first when no exact state-level match could be identified.

While PACER was demonstrated here using BC in the *AoU*, its underlying framework is broadly applicable to other diseases and biobank resources that integrate EHR, genomic, and survey-based data. Several limitations are worth noting. The quality of PACER-defined cohorts is inherently dependent on the completeness and consistency of EHR data, which can vary across *AoU* participants, and contributing health systems. For example, International Classification of Diseases 9^th^ and 10^th^ Revision (ICD-9/ICD-10) diagnostic codes are primarily assigned for administrative purposes, such as facilitating imaging authorization or billing, rather than reflecting a confirmed diagnosis (28). In addition, historical diagnoses made at external institutions may not be captured in the available records. Although requiring 2 independent BC diagnostic OMOP concept IDs reduces the likelihood of false-positive classification, true cases lacking sufficient documentation may be omitted or misclassified as controls (29,30). Additionally, while the matching strategy employed here accounts for key demographic and genetic ancestry factors, residual confounding from unmeasured variables cannot be fully excluded. Precise determination of the date of diagnosis remains challenging, as detailed pathology reports and operative notes are not available within the *AoU* CDR (version 8.0). As a result, analyses that depend on accurate time-to-event information, such as survival or interval-screening studies, should be interpreted with caution (31). The absence of tumor molecular profiling further limits our ability to stratify cases by subtype or to investigate subtype-specific risk factors in greater detail. In addition to EHR-derived factors, our study incorporates participants’ self-reported survey data to capture lifestyle, environmental, and socioeconomic factors. While these surveys provide valuable information independent of EHR, they introduce additional bias. Response rates vary across participants, and non-response may introduce systematic missingness that correlates with socioeconomic or other underlying factors (32). These limitations should be considered when interpreting associations involving factors generated from surveys.

## Conclusion

We present PACER, a customizable and reproducible phenotyping framework for constructing case and matched control cohorts within the *AoU*. By combining flexible EHR-based phenotyping with demographic and genetic ancestry matching, the framework supports more reliable investigation of disease risk across multiple data types. Using BC as a demonstration example, we combined EHR-based case phenotyping with a matched control algorithm incorporating sex at birth, age, predicted genetic ancestry, and state-level residency. We validated the resulting cohorts through concordance analyses across three independent approaches: phecodeX-defined cohorts, self-reported survey data, and genomic risk analyses, each providing consistent support for the quality of PACER-defined cohorts. Together, these findings demonstrate that PACER can identify well-defined case cohorts and generate well-matched controls that support integrated evaluation of clinical, genomic, lifestyle, and socioeconomic contributors to disease risk. PACER is a practical and reproducible framework for constructing well-defined case and matched control cohorts, with direct utility for studying disease risk, health disparities and strategies for early detection and prevention. As the *AoU* Research Program and similar large-scale biobanks continue to expand their participant diversity and data richness, frameworks such as PACER will be increasingly valuable for enabling systematic and reproducible investigation of multifactorial diseases across diverse populations.

## Acknowledgements

We gratefully acknowledge *All of Us* participants for their contributions, without whom this research would not have been possible. We also thank the National Institutes of Health’s *All of Us* Research Program for making available the participant data examined in this study.

## Author contributions

Yuewen Qi (Conceptualization, Data curation, Formal analysis, Methodology, Validation, Visualization, Writing—original draft, review & editing), Kassidy Lundy-Perez (Formal analysis, Validation, Visualization, Writing—original draft, review & editing), Devin Gee (Formal analysis, Validation, Visualization, Writing— original draft, review & editing), and Nyasha Chambwe (Conceptualization, Funding acquisition, Project administration, Supervision, Validation, Writing—review & editing).

## Data availability

The data analyzed in this study are available through the All of Us Researcher Workbench and are subject to the All of Us Research Program’s data access requirements and policies. The pipeline will be released on the All of Us Workbench as a community workspace resource for other users. The code used in this study is publicly available on GitHub at https://github.com/ChambweLab/PACER_All_of_Us and archived on Zenodo at https://doi.org/10.5281/zenodo.21829117.

## References

1. Sudlow C, Gallacher J, Allen N, Beral V, Burton P, Danesh J, et al. UK biobank: an open access resource for identifying the causes of a wide range of complex diseases of middle and old age. PLoS Med. 2015 Mar;12(3):e1001779. doi:10.1371/journal.pmed.1001779 PubMed PMID: 25826379; PubMed Central PMCID: PMC4380465.

2. All of Us Research Program Investigators, Denny JC, Rutter JL, Goldstein DB, Philippakis A, Smoller JW, et al. The “All of Us” Research Program. N Engl J Med. 2019 Aug 15;381(7):668–76. doi:10.1056/NEJMsr1809937 PubMed PMID: 31412182; PubMed Central PMCID: PMC8291101.

3. Bianchi DW, Brennan PF, Chiang MF, Criswell LA, D’Souza RN, Gibbons GH, et al. The All of Us research program is an opportunity to enhance the diversity of US biomedical research. Nat Med. 2024 Feb;30(2):330–3. doi:10.1038/s41591-023-02744-3 PubMed PMID: 38374344; PubMed Central PMCID: PMC11835384.

4. Ramirez AH, Sulieman L, Schlueter DJ, Halvorson A, Qian J, Ratsimbazafy F, et al. The All of Us Research Program: Data quality, utility, and diversity. Patterns. 2022 Aug 12;3(8):100570. doi:10.1016/j.patter.2022.100570 PubMed PMID: 36033590; PubMed Central PMCID: PMC9403360.

5. Getzen E, Ungar L, Mowery D, Jiang X, Long Q. Mining for equitable health: Assessing the impact of missing data in electronic health records. J Biomed Inform. 2023 Mar;139:104269. doi:10.1016/j.jbi.2022.104269 PubMed PMID: 36621750; PubMed Central PMCID: PMC10391553.

6. Kim MK, Rouphael C, McMichael J, Welch N, Dasarathy S. Challenges in and Opportunities for Electronic Health Record-Based Data Analysis and Interpretation. Gut Liver. 2024 Mar 15;18(2):201–8. doi:10.5009/gnl230272 PubMed PMID: 37905424; PubMed Central PMCID: PMC10938158.

7. Acosta JN, Leasure AC, Both CP, Szejko N, Brown S, Torres-Lopez V, et al. Cardiovascular Health Disparities in Racial and Other Underrepresented Groups: Initial Results From the All of Us Research Program. J Am Heart Assoc. 2021 Sep 7;10(17):e021724. doi:10.1161/JAHA.121.021724 PubMed PMID: 34431358; PubMed Central PMCID: PMC8649271.

8. Baierl J, Hsiao YW, Jones MR, Peng PC, Pharoah PDP. Measuring the accuracy of electronic health record-based phenotyping in the All of Us Research Program to optimize statistical power for genetic association testing. J Am Med Inform Assoc JAMIA. 2026 Mar 1;33(3):611–20. doi:10.1093/jamia/ocaf234 PubMed PMID: 41528460; PubMed Central PMCID: PMC12981652.

9. Zeng C, Schlueter DJ, Tran TC, Babbar A, Cassini T, Bastarache LA, et al. Comparison of phenomic profiles in the All of Us Research Program against the US general population and the UK Biobank. J Am Med Inform Assoc JAMIA. 2024 Jan 23;31(4):846–54. doi:10.1093/jamia/ocad260 PubMed PMID: 38263490; PubMed Central PMCID: PMC10990551.

10. Aschebrook-Kilfoy B, Zakin P, Craver A, Shah S, Kibriya MG, Stepniak E, et al. An Overview of Cancer in the First 315,000 All of Us Participants. PLoS ONE. 2022 Sep 1;17(9):e0272522. doi:10.1371/journal.pone.0272522 PubMed PMID: 36048778; PubMed Central PMCID: PMC9436122.

11. Bastarache L. Using Phecodes for Research with the Electronic Health Record: From PheWAS to PheRS. Annu Rev Biomed Data Sci. 2021 Jul 20;4(Volume 4, 2021):1–19. doi:10.1146/annurev-biodatasci-122320-112352

12. Shuey MM, Stead WW, Aka I, Barnado AL, Bastarache JA, Brokamp E, et al. Next-generation phenotyping: introducing phecodeX for enhanced discovery research in medical phenomics. Bioinformatics. 2023 Nov 1;39(11):btad655. doi:10.1093/bioinformatics/btad655 PubMed PMID: 37930895; PubMed Central PMCID: PMC10627409.

13. Kirby JC, Speltz P, Rasmussen LV, Basford M, Gottesman O, Peissig PL, et al. PheKB: a catalog and workflow for creating electronic phenotype algorithms for transportability. J Am Med Inform Assoc JAMIA. 2016 Nov;23(6):1046–52. doi:10.1093/jamia/ocv202 PubMed PMID: 27026615; PubMed Central PMCID: PMC5070514.

14. Iwagami M, Shinozaki T. Introduction to Matching in Case-Control and Cohort Studies. Ann Clin Epidemiol. 2022 Apr 4;4(2):33–40. doi:10.37737/ace.22005 PubMed PMID: 38504854; PubMed Central PMCID: PMC10760465.

15. Łukasiewicz S, Czeczelewski M, Forma A, Baj J, Sitarz R, Stanisławek A. Breast Cancer— Epidemiology, Risk Factors, Classification, Prognostic Markers, and Current Treatment Strategies— An Updated Review. Cancers. 2021 Aug 25;13(17):4287. doi:10.3390/cancers13174287 PubMed PMID: 34503097; PubMed Central PMCID: PMC8428369.

16. Jatoi I, Sung H, Jemal A. The Emergence of the Racial Disparity in U.S. Breast-Cancer Mortality. N Engl J Med. 2022 Jun 23;386(25):2349–52. doi:10.1056/NEJMp2200244 PubMed PMID: 35713541.

17. Breast Cancer | PheKB [Internet]. [cited 2026 Apr 21]. Available from: https://phekb.org/phenotype/breast-cancer

18. Chang CC, Chow CC, Tellier LC, Vattikuti S, Purcell SM, Lee JJ. Second-generation PLINK: rising to the challenge of larger and richer datasets. GigaScience. 2015;4:7. doi:10.1186/s13742-015-0047-8 PubMed PMID: 25722852; PubMed Central PMCID: PMC4342193.

19. Sharma S, Nagar SD, Pemu P, Zuchner S, SEEC Consortium, Mariño-Ramírez L, et al. Genetic ancestry and population structure in the All of Us Research Program cohort. Nat Commun. 2025 May 3;16(1):4123. doi:10.1038/s41467-025-59351-8 PubMed PMID: 40319026; PubMed Central PMCID: PMC12049439.

20. Hu J, Ye Y, Zhou G, Zhao H. Using clinical and genetic risk factors for risk prediction of 8 cancers in the UK Biobank. JNCI Cancer Spectr. 2024 Feb 29;8(2):pkae008. doi:10.1093/jncics/pkae008 PubMed PMID: 38366150; PubMed Central PMCID: PMC10919929.

21. Collister JA, Liu X, Clifton L. Calculating Polygenic Risk Scores (PRS) in UK Biobank: A Practical Guide for Epidemiologists. Front Genet. 2022;13:818574. doi:10.3389/fgene.2022.818574 PubMed PMID: 35251129; PubMed Central PMCID: PMC8894758.

22. Tzenios N, Tazanios ME, Chahine M. The impact of BMI on breast cancer – an updated systematic review and meta-analysis. Medicine (Baltimore). 2023 Feb 2;103(5):e36831. doi:10.1097/MD.0000000000036831 PubMed PMID: 38306546; PubMed Central PMCID: PMC10843423.

23. McGuire A, Brown JAL, Malone C, McLaughlin R, Kerin MJ. Effects of Age on the Detection and Management of Breast Cancer. Cancers. 2015 May 22;7(2):908–29. doi:10.3390/cancers7020815 PubMed PMID: 26010605; PubMed Central PMCID: PMC4491690.

24. Cerezo M, Sollis E, Ji Y, Lewis E, Abid A, Bircan KO, et al. The NHGRI-EBI GWAS Catalog: standards for reusability, sustainability and diversity. Nucleic Acids Res. 2025 Jan 6;53(D1):D998–1005. doi:10.1093/nar/gkae1070 PubMed PMID: 39530240; PubMed Central PMCID: PMC11701593.

25. Fatumo S, Chikowore T, Choudhury A, Ayub M, Martin AR, Kuchenbaecker K. A roadmap to increase diversity in genomic studies. Nat Med. 2022 Feb;28(2):243–50. doi:10.1038/s41591-021-01672-4 PubMed PMID: 35145307; PubMed Central PMCID: PMC7614889.

26. Huang H, Couch RE, Karam R, Hu C, Boddicker N, Polley EC, et al. Pathogenic Variants in Cancer Susceptibility Genes Predispose to Ductal Carcinoma In Situ of the Breast. Clin Cancer Res Off J Am Assoc Cancer Res. 2025 Jan 6;31(1):130–8. doi:10.1158/1078-0432.CCR-24-1884 PubMed PMID: 39513960; PubMed Central PMCID: PMC11701432.

27. Sondka Z, Bamford S, Cole CG, Ward SA, Dunham I, Forbes SA. The COSMIC Cancer Gene Census: describing genetic dysfunction across all human cancers. Nat Rev Cancer. 2018 Nov;18(11):696–705. doi:10.1038/s41568-018-0060-1 PubMed PMID: 30293088; PubMed Central PMCID: PMC6450507.

28. Holmes JH, Beinlich J, Boland MR, Bowles KH, Chen Y, Cook TS, et al. Why Is the Electronic Health Record So Challenging for Research and Clinical Care? Methods Inf Med. 2021 May;60(1– 02):32–48. doi:10.1055/s-0041-1731784 PubMed PMID: 34282602; PubMed Central PMCID: PMC9295893.

29. Ye Z, Mayer J, Ivacic L, Zhou Z, He M, Schrodi SJ, et al. Phenome-wide association studies (PheWASs) for functional variants. Eur J Hum Genet EJHG. 2015 Apr;23(4):523–9. doi:10.1038/ejhg.2014.123 PubMed PMID: 25074467; PubMed Central PMCID: PMC4666492.

30. Verma A, Ritchie MD. Current Scope and Challenges in Phenome-Wide Association Studies. Curr Epidemiol Rep. 2017 Dec;4(4):321–9. doi:10.1007/s40471-017-0127-7 PubMed PMID: 29545989; PubMed Central PMCID: PMC5846687.

31. Hersh WR, Weiner MG, Embi PJ, Logan JR, Payne PRO, Bernstam EV, et al. Caveats for the Use of Operational Electronic Health Record Data in Comparative Effectiveness Research. Med Care. 2013 Aug;51(8 0 3):S30–7. doi:10.1097/MLR.0b013e31829b1dbd PubMed PMID: 23774517; PubMed Central PMCID: PMC3748381.

32. Tesfaye S, Cronin RM, Lopez-Class M, Chen Q, Foster CS, Gu CA, et al. Measuring social determinants of health in the All of Us Research Program. Sci Rep. 2024 Apr 16;14(1):8815. doi:10.1038/s41598-024-57410-6

