## Supplemental pdf for "Rigorous Female Breast Cancer Phenotyping Using the *All of Us* Research Program"

### Supplemental Material

#### Supplemental Figures

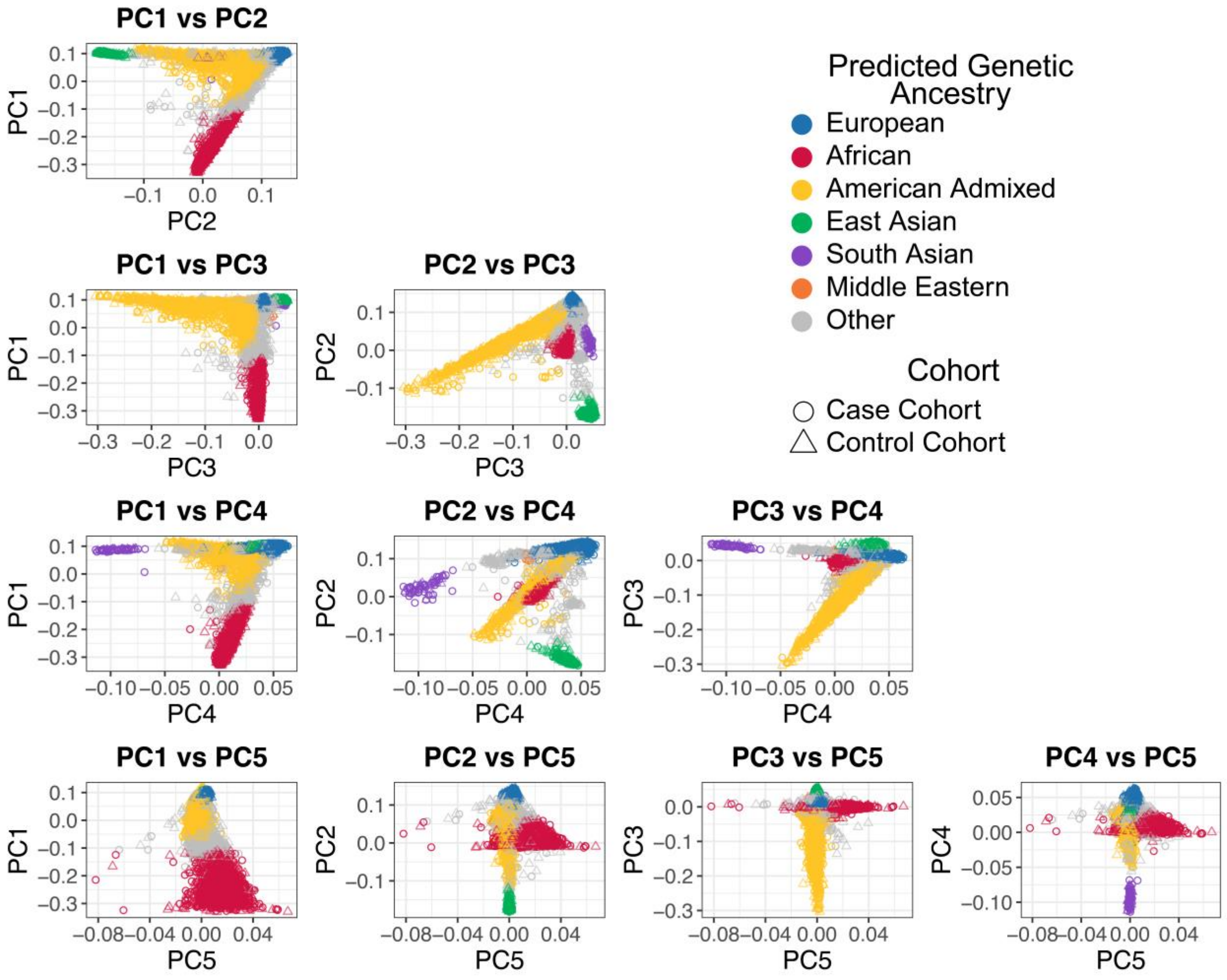


**Figure S1: Principal Component Analysis.** Analysis of *AoU* srWGS genotype calls in principal component space. Participants predicted genetic ancestry and case or control status are shown across combinations of the top 5 principal components.

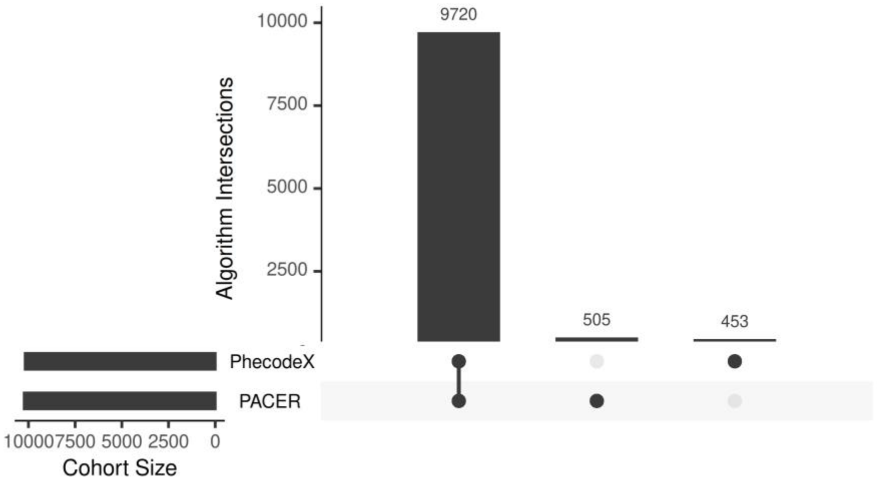


**Figure S2: Overlap between phecodeX and PACER breast cancer case cohorts.** The number of overlapping and differing participants between the BC cohort generated by phecodeX or PACER is displayed. PhecodeX was set to require 2 relevant BC International Classification of Disease identification codes at least 30 days apart.

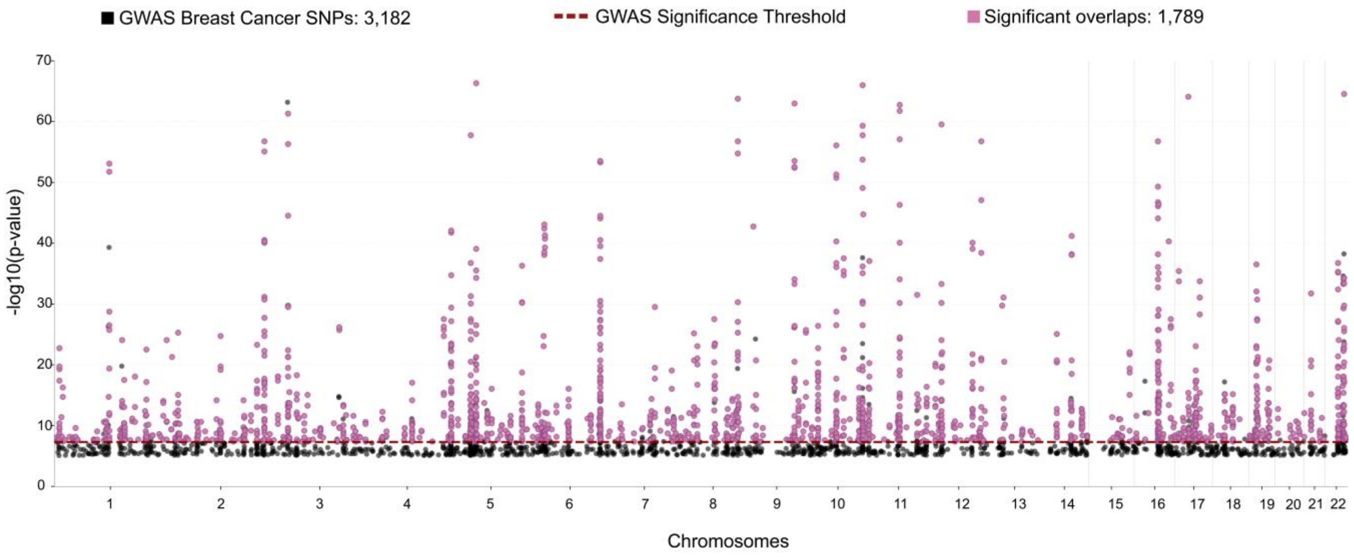


**Figure S3: Enrichment of GWAS catalog breast cancer SNPs in PACER breast cancer cohort.** The positional overlaps between SNPs identified by the GWAS catalog as significantly associated with BC traits, which were tested for enrichment within BC cohort genomes with respect to GWAS significance.

#### Supplemental Tables

| Name | OMOP Concept ID | SNOMED Code |
|---|---|---|
| Adenocarcinoma of breast | 3655521 | 865954003 |
| Carcinoma in situ of female breast | 4242800 | 92593006 |
| Carcinoma of breast | 4116071 | 254838004 |
| Carcinoma of breast - lower, inner quadrant | 4113637 | 286894008 |
| Carcinoma of breast - upper, inner quadrant | 4117850 | 286893002 |
| Carcinoma of breast - upper, outer quadrant | 4117851 | 286895009 |
| Carcinoma of central portion of breast | 46270923 | 708921005 |
| Carcinoma of female breast | 40486563 | 447782002 |
| Carcinoma of male breast | 4157448 | 372096000 |
| HER2-positive carcinoma of breast | 4142116 | 427685000 |
| Hormone receptor positive malignant neoplasm of breast | 4216891 | 417181009 |
| Human epidermal growth factor 2 negative carcinoma of breast | 4330242 | 431396003 |
| Infiltrating duct carcinoma of breast | 4237178 | 408643008 |
| Infiltrating duct carcinoma of female breast | 40492507 | 448952004 |
| Infiltrating duct carcinoma of left female breast | 759932 | 1080101000119100 |
| Infiltrating duct carcinoma of right female breast | 759933 | 1080181000119100 |
| Infiltrating ductal carcinoma of upper outer quadrant of right female breast | 36712723 | 1080241000119100 |
| Infiltrating lobular carcinoma of breast | 4080865 | 278054005 |
| Infiltrating lobular carcinoma of left female breast | 36712724 | 1080261000119100 |
| Infiltrating lobular carcinoma of right female breast | 36712725 | 1080341000119100 |
| Inflammatory carcinoma of breast | 4112074 | 254840009 |
| Intraductal carcinoma in situ of bilateral breasts | 761183 | 15636951000119100 |
| Intraductal carcinoma in situ of breast | 4001670 | 109889007 |

| | | |
|---|---|---|
| Intraductal carcinoma in situ of left breast | 759930 | 1079811000119100 |
| Intraductal carcinoma in situ of right breast | 759931 | 1079821000119110 |
| Invasive carcinoma of breast | 37017351 | 713609000 |
| Local recurrence of malignant tumor of breast | 4201477 | 314955001 |
| Malignant neoplasm of axillary tail of breast | 4155292 | 372094002 |
| Malignant neoplasm of axillary tail of female breast | 4091467 | 188156001 |
| Malignant neoplasm of breast lower inner quadrant | 4188544 | 373080008 |
| Malignant neoplasm of breast lower outer quadrant | 4187848 | 373081007 |
| Malignant neoplasm of breast upper inner quadrant | 4187849 | 373082000 |
| Malignant neoplasm of breast upper outer quadrant | 4160780 | 373083005 |
| Malignant neoplasm of central part of female breast | 4092511 | 188151006 |
| Malignant neoplasm of ectopic site of male breast | 4091471 | 188168005 |
| Malignant neoplasm of female breast | 4157332 | 372064008 |
| Malignant neoplasm of lower-inner quadrant of female breast | 4095740 | 188153009 |
| Malignant neoplasm of lower-outer quadrant of female breast | 4091466 | 188155002 |
| Malignant neoplasm of male breast | 4157447 | 372095001 |
| Malignant neoplasm of nipple and areola of female breast | 4091464 | 188147009 |
| Malignant neoplasm of nipple and areola of male breast | 4091469 | 188163001 |
| Malignant neoplasm of upper-inner quadrant of female breast | 4092512 | 188152004 |
| Malignant neoplasm of upper-outer quadrant of female breast | 4091465 | 188154003 |
| Malignant neoplasm, overlapping lesion of breast | 4092513 | 188157005 |
| Malignant phyllodes tumor of breast | 4112854 | 254844000 |
| Metastatic human epidermal growth factor 2 positive carcinoma of breast | 36684948 | 459391000124109 |
| Mucinous carcinoma of breast | 40480651 | 444712000 |
| Overlapping malignant neoplasm of female breast | 133711 | 109886000 |
| Overlapping malignant neoplasm of male breast | 4003684 | 109887009 |
| Paget's disease of nipple | 4301516 | 403946000 |

| | | |
|---|---|---|
| Papillary carcinoma in situ of breast | 36715826 | 721594000 |
| Primary malignant neoplasm of areola of female breast | 433148 | 93680004 |
| Primary malignant neoplasm of axillary tail of breast | 441513 | 372092003 |
| Primary malignant neoplasm of axillary tail of left female breast | 36684817 | 353421000119109 |
| Primary malignant neoplasm of axillary tail of right female breast | 36684820 | 353501000119104 |
| Primary malignant neoplasm of breast | 4162253 | 372137005 |
| Primary malignant neoplasm of breast lower inner quadrant | 4187851 | 373090000 |
| Primary malignant neoplasm of breast lower outer quadrant | 4188545 | 373091001 |
| Primary malignant neoplasm of breast upper inner quadrant | 4158563 | 373089009 |
| Primary malignant neoplasm of breast upper outer quadrant | 4187850 | 373088001 |
| Primary malignant neoplasm of central portion of female breast | 432845 | 93745008 |
| Primary malignant neoplasm of female breast | 137809 | 93796005 |
| Primary malignant neoplasm of female left breast | 36684818 | 353431000119107 |
| Primary malignant neoplasm of female right breast | 765123 | 353511000119101 |
| Primary malignant neoplasm of lower inner quadrant of female breast | 432263 | 93874009 |
| Primary malignant neoplasm of lower outer quadrant of female breast | 441515 | 93876006 |
| Primary malignant neoplasm of male breast | 135489 | 93884005 |
| Primary malignant neoplasm of skin of breast | 4247348 | 94012007 |
| Primary malignant neoplasm of upper inner quadrant of female breast | 440956 | 94115006 |
| Primary malignant neoplasm of upper outer quadrant of female breast | 436353 | 94117003 |
| Recurrent primary malignant neoplasm of left female breast | 36712738 | 1081551000119110 |
| Sarcoma of breast | 4175531 | 278050001 |
| Sarcoma of female breast | 40489942 | 448449001 |
| Secondary malignant neoplasm of breast | 603292 | 145501000119108 |
| Secondary malignant neoplasm of female breast | 140960 | 94297009 |
| Triple-negative breast cancer | 45768522 | 706970001 |

**Table S1: BC diagnostic OMOP concept IDs.** List of BC diagnostic OMOP concept IDs in AoU v8, including name and Systematized Nomenclature of Medicine (SNOMED) codes.

| Name | OMOP Concept Id | SNOMED Code |
|---|---|---|
| Estrogen receptor positive tumor | 4167696 | 416053008 |
| Hormone receptor positive malignant neoplasm of breast | 4216891 | 417181009 |
| Hormone receptor positive tumor | 4167363 | 417742002 |
| Progesterone receptor positive tumor | 4219694 | 416561008 |

**Table S2: HR+, ER+, and PR+ BC codes.** List of diagnostic OMOP concept IDs for HR+ breast cancer. Any BC case with at least one OMOP concept IDs in this table is classified as an HR+ BC case.

| Name | OMOP Concept ID | RxNorm Code |
|---|---|---|
| Anastrozole | 1348265 | 84857 |
| Exemestane | 1398399 | 258494 |
| Fulvestrant | 1304044 | 282357 |
| Goserelin | 1366310 | 50610 |
| Letrozole | 1315946 | 72965 |
| Leuprolide | 1351541 | 42375 |
| Tamoxifen | 1436678 | 10324 |
| Toremifene | 1342346 | 38409 |

**Table S3: Hormone drug OMOP concept IDs.** List of hormone drug usage OMOP concept IDs for identifying HR+ BC cases. BC cases with at least one hormone drug OMOP concept IDs recorded after their BC diagnostic OMOP concept IDs (**Table S1**) are classified as HR+ BC cases.

| Name | OMOP Concept Id | SNOMED Code |
|---|---|---|
| Abnormal findings on diagnostic imaging of breast | 434169 | 274530001 |
| Abscess of breast | 4153106 | 28432003 |
| Absence of breast | 4088290 | 248802009 |
| Acquired absence of breast | 36715792 | 721551005 |
| Adenocarcinoma of breast | 3655521 | 865954003 |
| At increased risk of malignant neoplasm of breast | 3655725 | 866242004 |
| Atypical lobular hyperplasia of left breast | 37209139 | 10836201000119100 |
| Benign mammary dysplasia | 78200 | 57993004 |
| Benign neoplasm of left breast | 37208024 | 350821000119101 |
| Benign neoplasm of right breast | 37208028 | 351381000119101 |
| Benign tumor of breast | 72576 | 269485000 |
| Breast lump | 80767 | 89164003 |
| Breast signs and symptoms | 4056770 | 198116001 |
| Calcification of breast | 4205375 | 309587003 |
| Carcinoma in situ of breast | 81250 | 189336000 |
| Carcinoma in situ of female breast | 4242800 | 92593006 |
| Carcinoma in situ of left breast | 601142 | 353591000119105 |
| Carcinoma of breast | 4116071 | 254838004 |
| Carcinoma of breast - lower, inner quadrant | 4113637 | 286894008 |
| Carcinoma of breast - upper, inner quadrant | 4117850 | 286893002 |
| Carcinoma of breast - upper, outer quadrant | 4117851 | 286895009 |
| Carcinoma of central portion of breast | 46270923 | 708921005 |
| Carcinoma of female breast | 40486563 | 447782002 |
| Carcinoma of male breast | 4157448 | 372096000 |
| Cyst of breast | 4161410 | 399294002 |
| Deformity of reconstructed breast | 45757371 | 123591000119103 |
| Disorder of breast | 77030 | 79604008 |
| Fibroadenosis of breast | 75010 | 23260002 |
| Fibrocystic disease of breast | 78804 | 27431007 |
| HER2-positive carcinoma of breast | 4142116 | 427685000 |
| Hormone receptor positive malignant neoplasm of breast | 4216891 | 417181009 |
| Human epidermal growth factor 2 negative carcinoma of breast | 4330242 | 431396003 |
| Hypertrophy of breast | 78474 | 372281005 |
| Infiltrating duct carcinoma of breast | 4237178 | 408643008 |
| Infiltrating duct carcinoma of female breast | 40492507 | 448952004 |
| Infiltrating duct carcinoma of left female breast | 759932 | 1080101000119100 |

| | | |
|---|---|---|
| Infiltrating duct carcinoma of right female breast | 759933 | 1080181000119100 |
| Infiltrating ductal carcinoma of upper outer quadrant of right female breast | 36712723 | 1080241000119100 |
| Infiltrating lobular carcinoma of breast | 4080865 | 278054005 |
| Infiltrating lobular carcinoma of left female breast | 36712724 | 1080261000119100 |
| Infiltrating lobular carcinoma of right female breast | 36712725 | 1080341000119100 |
| Inflammatory carcinoma of breast | 4112074 | 254840009 |
| Inflammatory disorder of breast | 79072 | 266579006 |
| Injury of breast | 4265765 | 62112002 |
| Intraductal carcinoma in situ of bilateral breasts | 761183 | 15636951000119100 |
| Intraductal carcinoma in situ of breast | 4001670 | 109889007 |
| Intraductal carcinoma in situ of left breast | 759930 | 1079811000119100 |
| Intraductal carcinoma in situ of right breast | 759931 | 1079821000119110 |
| Invasive carcinoma of breast | 37017351 | 713609000 |
| Lobular carcinoma in situ of breast | 4001315 | 109888004 |
| Local recurrence of malignant tumor of breast | 4201477 | 314955001 |
| Lump in left breast | 760895 | 12240181000119100 |
| Lump in right breast | 765053 | 12240221000119100 |
| Lump in upper outer quadrant of left breast | 764594 | 457191000124103 |
| Lump in upper outer quadrant of right breast | 764597 | 457231000124108 |
| Lump of lower inner quadrant of breast | 37311340 | 816053001 |
| Lump of lower outer quadrant of breast | 37311339 | 816054007 |
| Lump of upper inner quadrant of breast | 37311341 | 816052006 |
| Lump of upper outer quadrant of breast | 37311338 | 816055008 |
| Malignant neoplasm of axillary tail of breast | 4155292 | 372094002 |
| Malignant neoplasm of axillary tail of female breast | 4091467 | 188156001 |
| Malignant neoplasm of breast lower inner quadrant | 4188544 | 373080008 |
| Malignant neoplasm of breast lower outer quadrant | 4187848 | 373081007 |
| Malignant neoplasm of breast upper inner quadrant | 4187849 | 373082000 |
| Malignant neoplasm of breast upper outer quadrant | 4160780 | 373083005 |
| Malignant neoplasm of central part of female breast | 4092511 | 188151006 |
| Malignant neoplasm of ectopic site of male breast | 4091471 | 188168005 |
| Malignant neoplasm of female breast | 4157332 | 372064008 |
| Malignant neoplasm of lower-inner quadrant of female breast | 4095740 | 188153009 |
| Malignant neoplasm of lower-outer quadrant of female breast | 4091466 | 188155002 |
| Malignant neoplasm of male breast | 4157447 | 372095001 |
| Malignant neoplasm of nipple and areola of female breast | 4091464 | 188147009 |
| Malignant neoplasm of nipple and areola of male breast | 4091469 | 188163001 |
| Malignant neoplasm of upper-inner quadrant of female breast | 4092512 | 188152004 |
| Malignant neoplasm of upper-outer quadrant of female breast | 4091465 | 188154003 |

| | | |
|---|---|---|
| Malignant neoplasm, overlapping lesion of breast | 4092513 | 188157005 |
| Malignant phyllodes tumor of breast | 4112854 | 254844000 |
| Malignant tumor of breast | 4112853 | 254837009 |
| Mammographic calcification of breast | 44783760 | 697944008 |
| Mammographic microcalcification of breast | 45757639 | 27931000119107 |
| Metastatic human epidermal growth factor 2 positive carcinoma of breast | 36684948 | 459391000124109 |
| Microcalcifications of the breast | 72737 | 44771000 |
| Mucinous carcinoma of breast | 40480651 | 444712000 |
| Multiple cysts of breast | 42709964 | 449836005 |
| Neoplasm of breast | 81251 | 126926005 |
| Neoplasm of female breast | 4131014 | 126927001 |
| Neoplasm of upper outer quadrant of female breast | 4131763 | 126933005 |
| Overlapping malignant neoplasm of female breast | 133711 | 109886000 |
| Overlapping malignant neoplasm of male breast | 4003684 | 109887009 |
| Paget's disease of nipple | 4301516 | 403946000 |
| Pain of breast | 73819 | 53430007 |
| Papillary carcinoma in situ of breast | 36715826 | 721594000 |
| Primary malignant neoplasm of areola of female breast | 433148 | 93680004 |
| Primary malignant neoplasm of axillary tail of breast | 441513 | 372092003 |
| Primary malignant neoplasm of axillary tail of left female breast | 36684817 | 353421000119109 |
| Primary malignant neoplasm of axillary tail of right female breast | 36684820 | 353501000119104 |
| Primary malignant neoplasm of breast | 4162253 | 372137005 |
| Primary malignant neoplasm of breast lower inner quadrant | 4187851 | 373090000 |
| Primary malignant neoplasm of breast lower outer quadrant | 4188545 | 373091001 |
| Primary malignant neoplasm of breast upper inner quadrant | 4158563 | 373089009 |
| Primary malignant neoplasm of breast upper outer quadrant | 4187850 | 373088001 |
| Primary malignant neoplasm of central portion of female breast | 432845 | 93745008 |
| Primary malignant neoplasm of female breast | 137809 | 93796005 |
| Primary malignant neoplasm of female left breast | 36684818 | 353431000119107 |
| Primary malignant neoplasm of female right breast | 765123 | 353511000119101 |
| Primary malignant neoplasm of lower inner quadrant of female breast | 432263 | 93874009 |
| Primary malignant neoplasm of lower outer quadrant of female breast | 441515 | 93876006 |
| Primary malignant neoplasm of male breast | 135489 | 93884005 |
| Primary malignant neoplasm of skin of breast | 4247348 | 94012007 |
| Primary malignant neoplasm of upper inner quadrant of female breast | 440956 | 94115006 |
| Primary malignant neoplasm of upper outer quadrant of female breast | 436353 | 94117003 |
| Recurrent primary malignant neoplasm of left female breast | 36712738 | 1081551000119110 |
| Sarcoma of breast | 4175531 | 278050001 |
| Sarcoma of female breast | 40489942 | 448449001 |

| | | |
|---|---|---|
| Secondary malignant neoplasm of breast | 603292 | 145501000119108 |
| Secondary malignant neoplasm of female breast | 140960 | 94297009 |
| Solitary cyst of breast | 78473 | 266578003 |
| Triple-negative breast cancer | 45768522 | 706970001 |

**Table S4: BC-related OMOP concept IDs.** List of suggestive but not definitive BC-related diagnostic OMOP concept IDs. Participants with these codes are excluded from the control pool. The table includes names, OMOP concept IDs, and SNOMED codes.

| Factors | Category | Case (n=10225) | HR+ Case (n=5386) |
|---|---|---|---|
| **Sex at Birth** | Female | 10225 (100.00) | 5386 (100.00) |
| **Age, y** | <40 | 114 (1.12) | 68 (1.27) |
| | 40–49 | 613 (6.00) | 343 (6.37) |
| | 50–59 | 1485 (14.52) | 804 (14.93) |
| | 60–64 | 1212 (11.85) | 671 (12.46) |
| | ≥65 | 6801 (66.51) | 3500 (64.98) |
| **Race** | White | 7040 (68.85) | 3800 (70.55) |
| | Black/Afr. Am. | 1254 (12.26) | 613 (11.38) |
| | Others/ Haw./Pac. Isl. | 1083 (10.59) | 539 (10.00) |
| | Multiracial | 312 (3.05) | 149 (2.77) |
| | Asian | 230 (2.25) | 125 (2.32) |
| | Am. Indian/Alaska Nat. | 63 (0.62) | 28 (0.52) |
| | Mid. East/N. Afr. | 55 (0.54) | 25 (0.46) |
| | Skipped | 188 (1.84) | 107 (1.99) |
| **Ethnicity** | Non-Hispanic | 8779 (85.86) | 4654 (86.41) |
| | Hispanic | 1166 (11.40) | 574 (10.66) |
| | Others | 92 (0.90) | 51 (0.95) |
| | Skipped | 188 (1.84) | 107 (1.99) |
| **Genomic Data Available (WGS)** | Yes | 8576 (83.87) | 4555 (84.57) |
| | No | 1649 (16.13) | 831 (15.43) |
| **Predicted Genetic Ancestry (P ≥ 0.75)** | AFR | 1162 (11.36) | 562 (10.43) |
| | AMR | 781 (7.64) | 366 (6.80) |
| | EAS/ SAS | 191 (1.87) | 112 (2.08) |
| | EUR | 5891 (57.61) | 3208 (59.56) |
| | MID/ OTH | 551 (5.39) | 307 (5.70) |
| | No Ancestry Data | 1649 (16.13) | 831 (15.43) |
| **BMI** | Underweight | 203 (1.99) | 95 (1.76) |
| | Normal weight | 2983 (29.17) | 1576 (29.26) |
| | Overweight | 3056 (29.89) | 1613 (29.95) |
| | Obesity | 3939 (38.52) | 2090 (38.80) |
| | No data | 44 (0.43) | 12 (0.22) |
| **Area Deprivation Index** | Mean | 0.313 | 0.311 |
| | Median | 0.302 | 0.298 |
| **Education Level** | Advanced degree | 2901 (28.37) | 1556 (28.89) |
| | College graduate | 2724 (26.64) | 1490 (27.66) |

| | | | |
|---|---|---|---|
| | College 1-3 | 2676 (26.17) | 1386 (25.73) |
| | 12 / GED | 1273 (12.45) | 660 (12.25) |
| | Grade 9–11 | 251 (2.45) | 114 (2.12) |
| | Grade 5–8 | 152 (1.49) | 72 (1.34) |
| | ≤Grade 4 | 89 (0.87) | 42 (0.78) |
| | Skipped | 159 (1.56) | 66 (1.23) |
| **Employment Status** | Retired | 4292 (41.98) | 2182 (40.51) |
| | Employed | 3114 (30.45) | 1774 (32.94) |
| | Unable to work | 986 (9.64) | 464 (8.61) |
| | Self-employed | 648 (6.34) | 340 (6.31) |
| | Homemaker | 517 (5.06) | 262 (4.86) |
| | Unemployed > 1 year | 308 (3.01) | 168 (3.12) |
| | Unemployed < 1 year | 147 (1.44) | 79 (1.47) |
| | Student/ Skipped | 213 (2.08) | 117 (2.17) |
| **Income** | >200k | 916 (8.96) | 513 (9.52) |
| | 150k-200k | 626 (6.12) | 379 (7.04) |
| | 100k-150k | 1286 (12.58) | 716 (13.29) |
| | 75k-100k | 1094 (10.70) | 616 (11.44) |
| | 50k-75k | 1248 (12.21) | 650 (12.07) |
| | 35k-50k | 849 (8.30) | 470 (8.73) |
| | 25k-35k | 683 (6.68) | 362 (6.72) |
| | 10k-25k | 977 (9.56) | 459 (8.52) |
| | <10k | 546 (5.34) | 255 (4.73) |
| | Skipped | 2000 (19.56) | 966 (17.94) |
| **Insurance (Yes/No)** | Yes | 9942 (97.23) | 5273 (97.90) |
| | No | 114 (1.11) | 39 (0.72) |
| | Do not know/ Skipped | 169 (1.66) | 74 (1.38) |
| **Insurance Type** | Medi GAP/ Medicare | 2180 (21.32) | 1083 (20.11) |
| | Employer/Union | 1763 (17.24) | 1000 (18.57) |
| | Medicaid | 661 (6.46) | 338 (6.28) |
| | Purchased | 505 (4.94) | 256 (4.75) |
| | Private | 430 (4.21) | 223 (4.14) |
| | Other health plan | 128 (1.25) | 63 (1.17) |
| | Military/ VA | 113 (1.10) | 48 (0.90) |
| | Single service/ SCHIP/Indian/No coverage/ Other government/ State sponsored | 77 (0.75) | 38 (0.71) |
| | Do not know/ Skipped | 4368 (42.72) | 2337 (43.39) |

| | | | |
|---|---|---|---|
| **Alcohol Use** | ≥4 per week | 1222 (11.95) | 646 (11.99) |
| | 2 to 3 per week | 1280 (12.52) | 690 (12.81) |
| | 2 to 4 per month | 1774 (17.35) | 978 (18.16) |
| | Monthly or less | 3100 (30.32) | 1642 (30.49) |
| | Never | 1829 (17.89) | 941 (17.47) |
| | Skipped | 118 (1.15) | 59 (1.10) |
| **Smoking Frequency** | Not at all | 3262 (31.90) | 1726 (32.05) |
| | Some day | 260 (2.54) | 136 (2.53) |
| | Every day | 408 (3.99) | 211 (3.92) |
| | Do not know/ Skipped | 46 (0.45) | 28 (0.52) |

**Table S5. Demographic, socioeconomic, and clinical summary for the *AoU* BC cases and BC HR+ cases.** Values are presented as numbers of participants with percentages in parentheses for the 10,225 cases and 5,386 HR+ cases.
